# Evidence for a moderating effect of drinking severity but not HIV status on brain age in heavy episodic drinkers

**DOI:** 10.1101/2021.04.03.21254870

**Authors:** Jonathan Ipser, John Joska, Tatum Sevenoaks, Hetta Gouse, Carla Freeman, Tobias Kaufmann, Ole A. Andreassen, Steve Shoptaw, Dan J. Stein

## Abstract

**Background:** Chronic HIV infection and alcohol use have been associated with brain changes and neurocognitive impairment. However, their combined effects are less well studied. We correlated measures of “brain age gap” (BAG) and neurocognitive impairment in participants with and without HIV/AIDS and heavy episodic drinking (HED). We predicted that BAG will be greater in PWH who engage in HED and that these effects will be particularly pronounced in frontoparietal and subcortical regions.

**Method:** 69 participants were recruited from a community health centre in Cape Town: HIV-/HED-(N = 17), HIV+/HED- (N = 14), HIV-/HED+ (N = 21), and HIV+/HED+ (N = 17). Brain age gap (BAG) was derived from thickness, area and volumetric measurements from the whole brain or one of six brain regions. Linear regression models were employed to identify differences in BAG between patient groups and controls. Associations between BAG and clinical indicators of HIV and HED status were also tested using bivariate statistical methods.

**Results:** Compared to controls, greater whole brain BAG was observed in HIV-/HED+ (Cohen’s d = 1.61, p < 0.001), and HIV+/HED+ (d = 1.52, p = 0.005) participants. Differences in BAG between patients and controls were observed subcortically, as for the cingulate and the parietal cortex. An exploratory analysis revealed that higher relative brain ageing in HED participants with the highest drinking scores (W = 66, p = 0.036) but did not vary as a function of nadir CD4 count or current HIV viral load.

**Conclusion:** The association between heavy episodic drinking and BAG, independent of HIV status, points to the importance of screening for and targeting alcohol use disorders in primary care. Our findings also point to the utility of assessing the contribution of brain regions to the BAG.

## 1. INTRODUCTION

The widespread availability of antiretroviral therapy (ART) has substantially increased longevity in People With HIV/AIDS (PWH).The overall proportion of patients with HIV-associated neurocognitive disorder (HAND) has remained at roughly 50% in both the pre-ART and post-ART eras, although severe forms are now far less common [1]. Older PWH display brain abnormalities and neurocognitive deficits that are greater than those observed in age-matched seronegative individuals and younger PWH [2, 3]. The clinical impact of age-linked neurocognitive impairment in PWH is likely to be considerable, with HAND linked to greater cost, length of hospital stay and mortality [4]. Heavy drinking is prevalent in PWH in LMIC, with an estimated one third (37%) of respondents surveyed across 8 HIV treatment clinics (N = 1503) in Cape Town reporting harmful and hazardous drinking patterns (Kader et al. 2014). Given that heavy drinking is a well-established predictor of structural and functional abnormalities () associated with senescence., it becomes of paramount importance to elucidate the disparate effects of heavy drinking on brain health in PWH in these settings (Shield et al. 2013).

Changes reminiscent of that seen with ageing are particularly apparent in the brains of PWH with HAND, at both the cellular (eg. presence of amyloid-beta and tau plaques), and macroscopic (e.g. volumetric brain changes) levels [5].

Within the last decade investigators have used computational methods such as machine learning (ML) to provide a brain-based biometric of age, in which a model is trained on a set of features extracted from a large training dataset of typically thousands of structural and/or functional MRI datasets, allowing for the model-based prediction of an individual’s brain age in independent data [10]. The most frequently employed outcome of interest is the difference between a person’s chronological age and their model-predicted brain age, referred heretofore as the brain age gap (BAG), with positive BAG values indicating inflated brain age estimates relative to chronological age. Positive BAG statistics have been consistently reported in the literature in patients diagnosed with a range of psychiatric and neurological disorders, with the most extreme values typically observed for the dementias (5-10 years) and more modest estimates for mood disorders and HIV [10–12]. BAG has also previously demonstrated utility as a predictor of neurocognitive function in both PWH and healthy controls[13, 14].

The handful of ML studies assessing the association of HIV with brain ageing [12, 14–16] have consistently reported a greater BAG in PWH compared to seronegative controls. Moreover, these studies have found evidence more consistent with an additive, rather than an accelerated effect of HIV on brain age. This is the case both in studies that have covaried duration of HIV infection with BAG [14], as well as in a study in a middle-aged cohort of PWH that employed a longitudinal study design to provide a more rigorous test of causality [15]. In contrast, in the only published study of brain age in alcohol dependent (AD) and non-dependent PWH, the majority of whom were assessed multiple times over a period ranging from 1 month to 8 years, a region of interest volumetric approach provided evidence for accelerated aging in the frontal and parietal cortex across all PWH regardless of drinking status [17].

Predicting the combined effect of HIV and heavy drinking on brain ageing is complicated by the fact that ML studies report differences between PWH and AD patients in the specific brain regions that are sensitive to ageing. For instance, Guha et al. [18] reported an apparent HIV-specific effect of age on the reduction of subcortical, but not cortical, volumes. This contrasts with the finding in Guggenmos et al. [19], in which an aging effect was observed in cortical and limbic but not in cerebellar and basal ganglia regions in AD patients. Nevertheless, alcohol dependence exacerbated HIV-associated reductions in the volumes of the cingulate, insula, parietal and temporal cortices in a study employing a region of interest analytic approach [17].

This study aims to test the hypotheses that both heavy episodic drinking (HED) and HIV will be associated with greater apparent ageing of the brain, with the largest BAG in those in whom these conditions are comorbid, and (b), and that differences between groups will differ by regions assessed, with subcortical, frontal and parietal cortices contributing most towards groups differences in BAG. The extent to which BAG is associated with clinical outcomes (drinking scores, HIV viral load, nadir CD4 counts) in patients was also assessed. It is hypothesised that both HIV infection and binge drinking will be associated with greater BAG, and that any synergistic effect of these disorders on brain age will be most apparent in brain regions that have previously been demonstrated as being vulnerable to HIV and/ or alcohol associated neurotoxicity, including the frontal and parietal cortices. Evidence for an interaction between age and the BAG in PWH relative to healthy controls would provide some support for the argument that HIV infection accelerates brain ageing, rather than merely accentuates it [10].

## 2. METHODS

### 2.1. Recruitment

Participants were recruited from a primary health-care clinic in the Khayelitsha township in the Cape Town metropole, as part of a pilot study to investigate the combined effect of HIV and binge drinking on frontal cortical brain function. Seronegative participants were recruited from the clinic’s voluntary counselling unit. All participants provided informed consent, and the study was approved by the University of Cape Town Human Research Ethics Committee (HREC 003/2015) and conducted according to the principles of the Helsinki Declaration [21].

Eligible participants were placed into one of 4 groups, with a target of 30 participants each, as determined by HIV and drinking status (HIV+HED+, HIV+HED-, HIV-HED+, HIV-HED-). Although no formal statistical matching was performed, participants were preferentially selected within HIV and HED strata to match on gender. Following screening procedures and neurocognitive testing (detailed below), right-handed participants were asked to come for a follow-up visit within two weeks of their initial visit, to undergo MRI scanning.

Initial screening for eligibility was conducted at the HIV treatment clinic. An appointment to attend a study visit at Groote Schuur Hospital was scheduled within two weeks of the initial screen for those individuals who were willing to participate in the study, and only after they had completed an informed consent form and met all inclusion criteria. Participant’s understanding and appreciation of any risks and benefits of the study was interrogated using the University of California, San Diego Brief Assessment of Capacity to Consent questionnaire (UBACC) [22]. On their arrival for the first study visit, they were given a breathalyser test to determine whether they have complied with instructions not to drink on the day of testing (and to rule out heavy drinking during the previous day). In the event of a positive breathalyser test result, the test visit was rescheduled for another day, for completion of the neurocognitive battery.

### 2.2. Study selection criteria

To be eligible for the study, all participants were required to be between the ages 18-65 (inclusive), and have completed at least seven years of formal education. All participants were required to screen negative for a history of severe mental illness, active major depression and a recent (12 month) substance abuse history for substances other than alcohol (assessed using the MINI, version 6 [23]). Cannabis use was allowed, on the condition that the participant did not use cannabis within 30 days prior to the screening session. Evidence for recent use of cannabis, as well as morphine, opioids, heroin, cocaine and methamphetamine was assessed by means of a urine drug panel. The provision of a urine sample also allowed for testing of cotinine, a metabolite of nicotine. Moderate to severe scores on the Beck Depression Inventory II (>=19) was exclusionary [24]. Central nervous system diseases such as Korsakoff’s syndrome and Wernicke’s encephalopathy, a history of head injury with loss of consciousness exceeding 30 minutes, current nicotine dependence or treatment with psychotropic medication, were all grounds for exclusion. Participants who were claustrophobic or who had metal objects in their heads were excluded from the imaging phase of the study. In order to minimize the effects of pregnancy-associated hormonal fluctuations on cognition and imaging, women of child-bearing age who indicated that they were pregnant were excluded from study participation.

Participants in the HIV+ groups were required to have a positive diagnosis of HIV infection using two independent rapid HIV test kits, with the ELISA test employed to reconcile discrepant test results. In addition, patients were restricted to those who were ART-naïve, or who had received ART for a period of at most one month at testing, in order to avoid confounding brain and neurocognitive sequelae of ART with those resulting from HIV infection or HED.

Participants in the HED+ groups were required to report drinking at least 6 standard drinks (a standard drink equals 14 grams of alcohol) per occasion on at least a weekly basis for the past year. Inclusion in the low/non drinking (HED-) groups was restricted to individuals who had at least one drink over the past year, but who did not drink more than 2 drinks on a typical day during that time period. Moreover, HED-subjects were also required to indicate that they drink at most 2 to 3 times a week.

### 2.3. Assessment of drinking severity amongst heavy episodic drinkers

Drinking severity was measured using a modified version of the Substance Abuse and Mental Illness Symptoms Screener (SAMISS), an instrument previously validated in a population of PWH in South Africa [25]. This 16 item Likert type scale contains 7 items assessing drinking behaviour, and 9 items assessing mental health symptoms. The first 3 items of the SAMISS correspond to items on the validated AUDIT-C drinking questionnaire [26], and elicit information about how often a person has had a drink containing alcohol (never, monthly or less, 2–4 times/mo, 2–3 times/wk, >=4 times/wk), how many drinks s/he has had on a typical day when drinking (none, 1 or 2, 3 or 4, 5 or 6, 7–9, >=10), and how often they have more than 3 drinks on one occasion (never, less than monthly, monthly, weekly, daily or almost daily).

### 2.4. Assessment of immunological status

HIV viral load and CD4 count were assayed from blood samples obtained at the test visit. For the minority of participants for whom this was not possible, routinely collected HIV viral load and CD4 count data from the clinic for the visit closest in time to the test session was employed.

### 2.5. Measurement of additional covariates

Body Mass Index (BMI) was calculated from height and weight measurements conducted in the clinic using regularly calibrated instruments. Childhood adversity was assessed using the total score from the 25 item Childhood Trauma Questionnaire (CTQ), with analysis including CTQ total score restricted to data judged as valid based on the 3 validation items included in this scale [35].

### 2.6. MRI acquisition

T1 MRI data was acquired using a multi-echo MPRAGE sequence (TR = 2000ms, TE = E[1,2,3,4] = 1.53ms, 3.21ms, 4.89ms, 6.57ms, flip angle = 7%, FoV = 256×256mm) optimized for tissue segmentation with Freesurfer [36]. Resting-state fMRI, magnetic resonance spectroscopy and diffusion tensor imaging data was acquired in the same scan session, but will not be considered further in this manuscript.

### 2.7. Data analysis

Bivariate associations between study group and demographic (eg. age, gender) and clinical (eg. current and nadir CD4 count, HIV viral load, drinking score) characteristics were tested using ANOVA for continuous variables, or the non-parametric equivalent in cases where assumptions were violated or group sample sizes were too small (N < 20) to test these assumptions. The same procedure was employed to test associations between BAG and these covariates. Confounders were identified as covariates that were significantly associated, at alpha < 0.1, with both group membership and BAG.

Differences in BAG for all patients versus controls were identified with linear regression models. Where group emerged as a significant predictor, an additional model in which the 4 study groups were dummy coded was tested to identify associations with specific patient samples. Regional specificity of ageing associations with HIV and binge drinking were assessed by training separate models on features extracted using the Freesurfer Lobestrict segmentation from occipital, frontal, temporal, parietal, cingulate insula and subcortical (including cerebellar) regions (see 11, Fig 1b).

**Figure 1.**
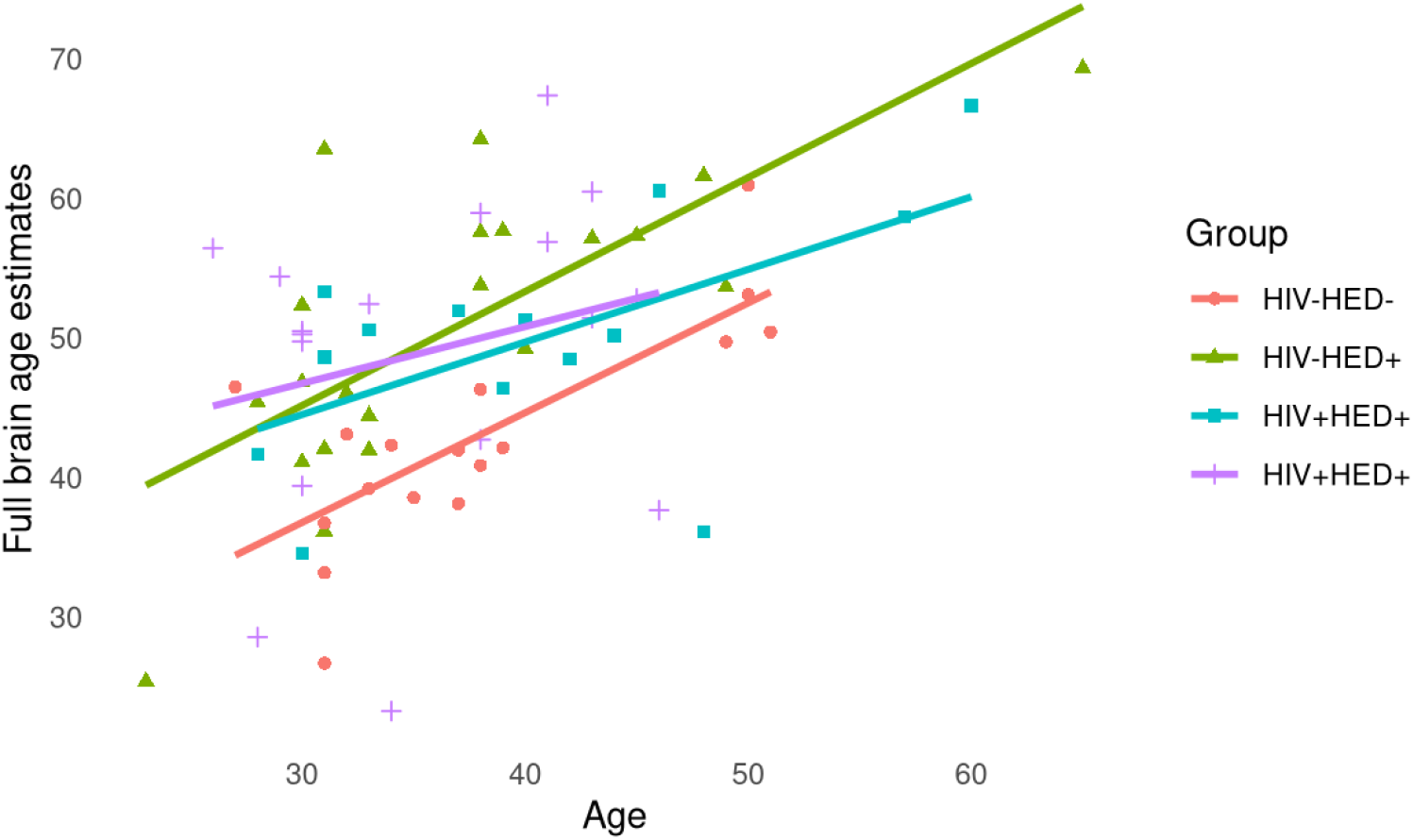
Correlation between chronological and brain age, by group

Age and the combined Euler statistic were included as covariates in all regression models. Age was included in these models to mitigate the statistically significant negative association observed between chronological age and BAG across the entire sample (R = −0.33, p < 0.01, see Fig. S2.), with the inclusion of the combined Euler statistic justified by its association with greater BAG (r = 0.37, p < 0.01). Sex was not included as a predictor given the use of sex-specific brain age models (see below), and evidence that sex was not significantly associated with either group status or BAG (see Table 1). All statistical analyses were conducted using R v4.0.2. [38].

**Table 1.** Demographic and clinical characteristics

|  | **HIV-HED-**<br>**(N = 17)** | **HIV-HED+**<br>**(N = 21)** | **HIV+HED-**<br>**(N = 14)** | **HIV+HED+**<br>**(N = 17)** | **Test** |
| --- | --- | --- | --- | --- | --- |
| **Age** | 37.82 (7.64) | 36.9 (9.36) | 40.43 (9.89) | 35.59 (6.63) | X<sup>2</sup> = 2.4, p= .49 |
| **Female** | 0.59 | 0.43 | 0.71 | 0.59 | X<sup>2</sup> = 2.93, p= .4 |
| **Education** | 10.47 (1.55) | 11 (1.49) | 10.15 (1.68) | 10.19 (1.8) | X<sup>2</sup> = 1.07, p= .78 |
| **Drinking score** | 4 (1.41) | 14.79 (1.78) | 4.42 (1.44) | 14.92 (1.44) | X<sup>2</sup> = 45.47, p< .001 |
| **BMI** | 30.06 (8.83) | 27.15 (8.34) | 25.15 (6.27) | 25.03 (7.07) | W = 1.41, p= .7 |
| **CTQ** | 50.46 (9.73) | 53.06 (14.15) | 53.17 (11.4) | 57 (12.88) | W = 2.42, p= .49 |
| **Nicotine+** | 0.24 | 0.52 | 0.21 | 0.59 | X<sup>2</sup> = 8.53, p= .036 |
| **HIV-load** |  |  | 8.59 (2.23) | 8.78 (1.81) | W = 64, p= .69 |
| **CD4-nadir** |  |  | 374.07 (197.16) | 394.27 (270.79) | W = 110, p= .85 |
| **Euler** | -36 (-67 to -25) | -43 (-88 to -21) | -23.5 (-31.5 to -21) | -28 (-63 to -21) | X<sup>2</sup> = 2.53, p= .47 |

Note: For continuous variables, such as years of education, results presented as mean; standard deviation. The log of the HIV viral load is presented in the table. Euler number is presented as the mean and interquartile range for averages of estimates for the left and right hemispheres

The clinical significance of accentuated brain age was assessed by regressing nadir CD4 cell counts and log-transformed HIV viral load from BAG, where applicable. In light of a possible ceiling effect on the drinking scale, a dichotomized version of the drinking score variable that distinguished between those achieving the maximal score and those who did not was included in analyses in non-parametric Mann-Whitney tests this study.

The methodology used in training the brain age model employed in this study has been described in detail elsewhere [11]. Briefly, a ML model based on gradient tree boosting and using the xgboost algorithm (as implemented in R [39]) was built for each sex (Males = 16,484; Females = 18,990) to predict the age of the brain from a set of thickness, area and volume features extracted using a multimodal parcellation of the cerebral cortex as well as a set of cerebellar–subcortical volume features (1,118 features in total). The training dataset for the model encompassed a wide age range (3-89 years), with a high degree of correspondence between chronological age and predicted brain age (*r* = 0.93 for the female model and *r* = 0.94 for the male model (11; Supplementary Fig. 2). These models were then applied using R scripts provided by 11 to segmented output from FreeSurfer 5.3 (https://surfer.nmr.mgh.harvard.edu), as derived from the T1-weighed MRI scans for the study participants. To validate the use of the training data model in relating brain age with chronological age for this paper, Pearson’s correlations (*R*), variance explained (*R*^*2*^), and mean absolute deviation (*MAD*) were calculated for the healthy control group in this study.

**Figure 2.**
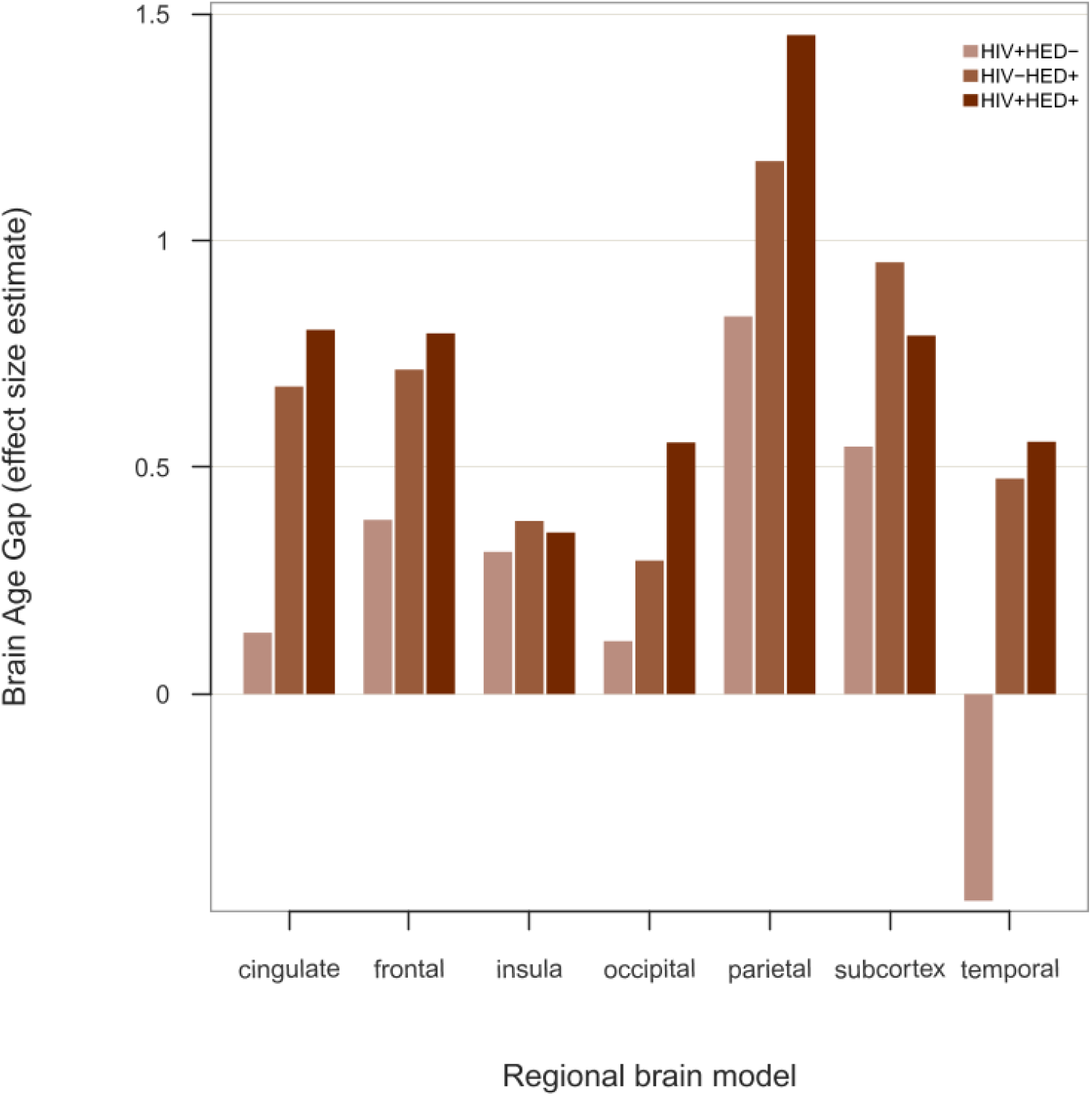
Regional differences in brain age gap by group

## 3. RESULTS

### 3.1. Sample description

A total of 69 participants were eligible for inclusion in the study. Non-parametric statistical tests revealed that groups were comparable on most demographic and clinical variables assessed, with the exception of drinking score and nicotine use (see Table 1). A moderately sized correlation between estimated brain age and chronological age was observed in the total sample (*R* = 0.53, p < .001), with the strongest correlation in the healthy control group (*R* = 0.75, p < .001) (see Fig. 1). The observation that over half of the individual variability in brain age in this particular group (*R*^*2*^ = 0.57) was accounted for by age (*MAD* = 4.04 years) provides some validation for using the Kaufmann et al. (2019) training model for this study. Visual assessment reveals the relationship between brain age estimates and chronological age to be roughly parallel for the study sub-groups, as illustrated in Fig. 1, and does not support an additive effect of HED on the brain in PWH, or indicate that chronic heavy drinking accelerated brain ageing in PWH.

Note: For continuous variables, such as years of education, results presented as mean; standard deviation. The log of the HIV viral load is presented in the table. Euler number is presented as the mean and interquartile range for averages of estimates for the left and right hemispheres

### 3.2. Identification of current nicotine use as a possible confound

As evident in Table 1, not only were total drinking scores significantly higher for the HED+ than HED-groups, as anticipated, but the former also tended to consist of a larger proportion of current smokers, as confirmed by cotinine urine assays (*X*^*2*^ = 8.53, *p* = 0.04). Nicotine use was identified as a potential confound of the association between HED and BAG, given that it was also associated with greater BAG (t = −2.58, p = 0.012). We therefore decided to include it as a covariate in all regression models. Although larger total CTQ scores were associated with greater BAG (*R* = 0.33, *p* = 0.012). and larger BMI predicted smaller BAG (*R* = −0.3, *p* = 0.041), these variables were not significantly different (at *p* < 0.1) between groups, and were not included as potential confounds in regression analyses. BAG did not vary as a function of years of education (*R* = 0.05, *p* = 0.686) or gender (*W* = 472, *p* = 0.174)

### 3.3. Greater BAG in binge drinkers, regardless of HIV serostatus

Confirmation that brain ageing is greater in the patient groups, relative to the healthy control participants, was obtained from the results of the regression model, adjusting for age, current nicotine use, and the Euler statistic (*β* = 6.47, *p* = 0.004). Inspection of pairwise contrasts revealed that while there was a numerically larger BAG for the HIV+/HED-than HIV-/HED-groups (mean = 9.52, SD = 8.54 versus mean = 5.13, SD = 5.47, Cohen’s *d* = 0.82), this difference was not statistically significant (*β* = 3.38, *p* = 0.22). In contrast, large and statistically significant effects were observed in the drinkers compared to the HIV-/HED-group, with similar brain age gaps observed for those who are seronegative (mean = 13.91, SD = 7.44, Cohen’s *d* = 1.61, *β* = 9.26, p < 0.001), as for those who were seropositive (mean = 13.44, SD = 11.79, Cohen’s *d* = 1.52, *β* = 7.53, *p* = 0.005). It is noteworthy that the BAG in the control group (HIV-/HED-) in this study was also significantly larger than zero (Wilcoxon signed rank *V* = 143, *p* = < 0.001).

### 3.4. Brain age gap varies by region of brain modelled

The proportion of variability in the whole brain BAG outcomes explained by regional models across the entire sample was greatest for the subcortex (*R*^*2*^ = 0.66), followed by the frontal cortex (*R*^*2*^ = 0.61). The insula and the occipital cortex contributed least to the predictive value of the full-brain model (*R*^*2*^ = 0.35 and *R*^*2*^ = 0.23, respectively). Estimates of variability explained by all the regional models can be found in the supplementary material. Differences in BAG were evident when comparing patients versus controls, after correcting for individual variability in age, current nicotine use, and Euler number, for the cingulate cortex (*β* = 5.14, *p* = 0.01), the subcortex (*β* = 9.02, *p* = 0.003), and the parietal cortex (*β* = 8.36, *p* = < 0.001). No evidence of greater brain ageing in patients was observed in the models trained on features from the occipital cortex (*β* = 1.81, *p* = 0.489), the insula (*β* = 2.51, *p* = 0.27), or the frontal cortex (*β* = 4.08, *p* = 0.07). Although these findings are partially consistent with our hypothesis that subcortical brain age estimates would be inflated in patients, where differences between patients and controls in regional BAG were observed, these differences were sensitive to drinking but not HIV status (see Table 2). Figure 2 provides a graphical representation of group differences in regional estimates of BAG.

**Table 2.** Regional BAG differences between patient groups and healthy controls

|  | HIV+HED- | HIV-HED+ | HIV+HED+ |
| --- | --- | --- | --- |
| **cingulate** | 2.135 (0.389) | 6.805 (0.004) | 6.389 (0.011) |
| **frontal** | 1.477 (0.607) | 5.974 (0.026) | 4.531 (0.113) |
| **insula** | 3.006 (0.31) | 3.408 (0.209) | 0.636 (0.827) |
| **occipital** | 1.437 (0.677) | 1.714 (0.587) | 2.397 (0.48) |
| **parietal** | 5.009 (0.065) | 9.314 (0.0) | 8.939 (0.001) |
| **subcortical** | 6.327 (0.112) | 11.365<br>(0.002) | 8.633 (0.029) |
| **temporal** | -3.320 (0.216) | 2.407 (0.325) | 0.962 (0.714) |

Note: Results presented as model coefficients, in units of years, for group membership relative to the HIV-HED-reference group (p-values in parentheses). Model adjusted for individual differences in age, current nicotine use, and average Euler number.

### 3.5. Greater relative brain age is linked to drinking severity but not HIV clinical variables

There was no evidence for an association between BAG and HIV viral load (*Rho* = −0.09, *p* = 0.682) or nadir CD4 cell count (*Rho* = −0.06, *p* = 0.770). Higher relative brain ageing was observed in those HED participants with the highest drinking scores (Mann-Whitney W = 66, p = 0.036). Notably, the effect size estimate for the relationship between BAG and drinking severity was similar in the HIV seropositive (*d* = 2.36) and seronegative (*d* = 2.13) drinkers.

## 4. DISCUSSION

In this study, a history of heavy episodic drinking, but not HIV status, was associated with greater BAG. HED over the last 3 months was associated with a large effect size of 1.57, indicating that in those with unhealthy drinking habits, BAG was greater than the mean for the control group in almost all participants (Cohen’s U_3_ = 94%). Moreover, participants with higher drinking scores displayed greater BAG, irrespective of HIV status.

Our data did not support an additive effect of HED on the brain in PWH, or indicate that chronic heavy drinking accelerated brain ageing in PWH. Studies testing which of the models of accentuated versus accelerated brain ageing best fit the data in cohorts of PWH have produced mixed results. A previously published association between age and subcortical volume and shape in PWH provides preliminary support for the accelerated ageing model [18, 40], and concurs with findings from a systematic review that 3 of 4 longitudinal studies documented accelerated decline in neurocognitive function in elderly PWH over time [41]. On the other hand, studies utilising ML have failed to uncover evidence of an interaction between HIV diagnosis and brain ageing, with findings supporting accentuated rather than accelerated effects of diagnosis (Cole et al. 2018; Cole et al. 2017; Kuhn et al. 2018). For instance, Cole et al [15] provided evidence supporting greater brain ageing in PWH versus controls, with BAG progressing at the same rate in both groups at 2 years follow-up.

Efforts to compare our findings with the published literature are complicated by the lack of comparable effect size statistics estimates for published brain age studies. Nevertheless, it is noteworthy that absolute BAG estimates in our HIV+ cohort are considerably higher than that reported by other brain age studies of PWH utilising ML algorithms, where equivalent statistics of 1 to 3 years have been reported in patients [14–16]. The same pattern is evident when comparing the HED sub-groups to data from age-equivalent alcohol dependent patients [19]. These findings, in conjunction with the large effect size for BAG for all clinical sub-groups is particularly striking given the relatively young cohort employed in this study, with an average age for the entire sample of 37 years (interquartile range: 30-43 years).

The inflated BAG estimates in this study may point to differences between the training dataset utilized for the sex-specific brain age models, comprised largely of well-educated individuals from well-resourced Northern hemisphere populations, and the test dataset, consisting of individuals from a relatively impoverished community in a developing nation. This is a possible explanation for the observation that the BAG in the HIV-/HED-control group was significantly larger than zero. Indeed, when one subtracts the mean BAG from the control group from that of the clinical groups, one observes group-specific BAG estimates that, although large, are more in line with the literature (mean: 4.47 (HIV+HED-); 8.79 (HIV-HED +); 8.32 (HIV+HED+)).

Nadir CD4 cell counts and current HIV viral load were not associated with BAG in our sample. The null finding with respect to nadir CD4 is consistent with findings from other studies of PWH using ML methods [14, 16]. Using white matter microstructural features to predict brain age, Kuhn et al. [12] did observe an interaction between chronological age, patient’s highest lifetime HIV RNA viral load and BAG, with greater BAG in older participants with higher self-reported lifetime viral load. Together, these findings suggest that neuroimaging modality and choice of HIV-related clinical variables are important factors to consider in planning research studies, as they may be informative with regards to brain age in PWH.

The finding that the largest associations between diagnosis and BAG were evident in heavy episodic drinkers is consistent with evidence that heavy episodic drinking (HED) or binge type drinking behaviour may have a negative impact on brain function [44]. Excess glutamate release associated with repeated cycles of alcohol consumption and withdrawal are liable to be particularly neurotoxic [45, 46]. Moreover, although not directly relevant to the findings of this study, ethanol may decrease intracellular exposure in HIV-infected monocyte cells to the ART drug, Elvitegravir [47], suggesting the possibility that PWH who engage in HED may not enjoy the benefits to brain health and neurocognitive function associated with adherence to a stable ART regimen.

Limitations of this study include the relatively small size of the subgroups, suggesting caution in interpreting the significance of models of ageing in this study [41]. Study participants were recruited shortly at the same clinic visit at which their HIV test results were disclosed by the clinic nurses. It was therefore not possible to test for evidence of accelerated brain age by relating BAG to duration of HIV infection. Nevertheless, there was no evidence for an interaction between age and HIV serostatus in this study.

There are a number of advantages to using BAG as an estimate of brain health in clinical studies. Firstly, the approach collapses a large amount of brain phenotypes into a single metric, which helps to minimise the risk of spurious false positive associations resulting from multiple comparisons, an endemic issue in neuroimaging studies. Second, it allows us to place brain images into a normative context. Plausibility of the BAG as a biomarker of relative brain health is also supported by the finding that brain age is to a large extent heritable, as reported in both GWAS and twin studies [11, 48]. Nevertheless, as a measure of brain age model accuracy, the BAG is a function of both the quality of data (noise) and true deviations from a normative brain developmental trajectory [11]. Although the average Euler number within groups compared favourably to that reported in the only other published study utilising the particular brain age model employed in this study [11], and was included in statistical models as a proxy measure of data quality, one cannot entirely discount poor signal to noise in the data as contributing to inflated BAG estimates in this study. Furthermore, it is worth noting that the model used in Kaufmann et al (2019) covered neurodevelopmental and old age spans better than the age span relevant to the predictions in our study. Thus, a model trained within this specific age range might in the future help further delineating the brain age effects.

Strengths of this study include the large training dataset employed, compared to other papers assessing brain ageing in PWH (eg. Cole et al. 2017), the use of a well-characterized clinical sample, the similar number of males and females in each of the comparison groups, and the exclusion of participants with diagnoses of comorbid depression. In addition, recruitment of ART-naïve PWH to investigate the association of substance abuse with brain health and neurocognitive function in PWH simplifies the interpretation of study findings, as one does not need to take possible effects of ART on the brain into account [9, 49].

Future investigations that evaluate the impact of SES factors (eg. access to resources, literacy, education) on brain age, are warranted. Adequately powered studies of the implications of greater brain age in patients groups on neurocognitive function and everyday functioning are also warranted. Research comparing the extent of convergence between brain age estimates using different algorithms, imaging modalities, and biomarkers of ageing, including telomere length and DNA methylation would also boost confidence in findings reported in the brain ageing literature. The observation of an interaction between age and HIV status in the shape of various subcortical regions (bilateral nucleus accumbens, amygdala, caudate, and thalamus as well as right pallidum and putamen) that was not detectable using standard volumetric measures for these regions suggest that it might be beneficial to train brain age models using these relatively novel metrics [40]. Finally, future investigations of mechanisms by which HIV and comorbid substance use might impact brain age, including mitochondrial DNA deletions [50], as well as possible hepatic dysfunction [17], are warranted.

## 5. CONCLUSION

Employing a factorial design, this study provided novel evidence for an association between heavy episodic drinking and greater relative brain age in both HIV seropositive and seronegative participants. The failure to detect evidence for accentuated or accelerated brain ageing in light/non-drinking PWH may be partly a function of the small sample of sub-groups, with resulting power constraints. Finally, the relatively large estimates of BAG in this study compared to those reported in both PWH and alcohol dependent cohorts in the extant literature could reflect the impoverished setting from which participants in this study were recruited, a possibility that warrants further investigation.

## Data Availability

The authors will exercise their duty to the study participants, as described in the patient informed consent forms, not to distribute the data to individuals who are not part of the investigative team publishing this study, given the sensitive nature of the data, and risks of stigmatisation associated with the possible identification of study participants should their data be distributed to other research groups.

## ^1^ Abbreviations

HED: heavy episodic drinking
BAG: brain age gap

## Author contributions

**Jonathan Ipser:** Conceptualization, Methodology, Formal analysis, Writing - Original Draft, Visualization, Project administration, Funding acquisition

**John Joska:** Conceptualization, Methodology, Writing - Review & Editing

**Tatum Sevenoaks:** Writing - Review & Editing

**Hetta Gouse:** Methodology, Investigation, Writing - Review & Editing

**Carla Freeman:** Investigation, Writing - Review & Editing

**Tobias Kaufmann:** Software, Writing - Review & Editing

**Ole A. Andreassen:** Writing - Review & Editing

**Steve Shoptaw:** Supervision, Writing - Review & Editing

**Dan J. Stein:** Supervision, Writing - Review & Editing

## Acknowledgements

Thanks go to Teboho Linda, who conducted all neurocognitive assessments for this study, Ntombekhaya Queen Maswana and Sheila Mdlakoma, who assisted with recruitment and participant screening, the Cape Universities Body Imaging Center (CUBIC) at Groote Schuur Hospital in Cape Town, where all the MRI scans were performed, and the study participants who gave so willingly of their time. Freesurfer 5.3 segmentation of the test dataset was performed using facilities provided by the University of Cape Town’s ICTS High Performance Computing team: hpc.uct.ac.za

